# Sc-RNA seq in familiar Gardner syndrome combined left atrial appendage fibroma reveals APC-C-MYC signaling modulates fibrotic subpopulation remodeling

**DOI:** 10.1101/2022.06.10.22274829

**Authors:** Xiaoping Li, Shengzhong Liu, Chenqing Zheng, Jichang Huang, Xun Xiao, Rong Luo, Hu Fan, Jiangtao Yang, Benqing Wu, Xiushan Wu, Wei Hua

## Abstract

**Background:** Gardner’s syndrome was once considered a subtype of familial adenomatous polyposis (FAP), and their molecular pathological features remain to be clarified. Familiar Gardner’s syndrome complicated by a rare giant left atrial appendage aneurysm (LAAA) is an unreported novel type of FAP syndrome, and exploring its causative cellular subtypes and molecular pathological features will provide new insights into the precise treatment of the syndrome.

**Methods:** Whole-exome sequencing was performed in the familial Gardner’s syndrome patients, and single cell sequencing of left atrial dilatation tumor, intestinal polyp and scalp cyst in the proband was performed to explore the cellular and molecular pathological mechanisms.

**Results:** Exon sequencing indicated the presence of a rare germline variant (c. 4666 dup A, p. Thr1556fs, rs587783031) in APC (the adenomatous polyposis coli gene), which caused changes in the APC related Wnt pathway. Sc-RNA seq in LAAA revealed an increased proportion of fibroblasts and immune cells. We re-clustered fibroblasts and identified five distinct sub-populations in LAAA: cancer associcated fibroblasts (CAF), cardiomyocyte like fibroblasts (CLF), endothelial like fibroblasts (ELF), T_cells like fibroblasts (TLF), and nomal like fibroblasts (NLF). Notably, nomal fibroblasts should constitute the main component of normal cardiac fibroblasts in LA tissues, while CAF mainly dominated in LAAA tissues. The trajectory and RNA velocity analysis revealed LAAA fibroblasts made a preferential transition from the immature phase (CLF, ELF, TLF, and NLF) at the apex of the trajectory to mature phase of tumor-like properties (CAF), indicating normal fibroblast was reprogrammed into CAF in LAAA. These results suggested diseased LA tissue with GS patient appears to display tumor-prone properties. Moreover, Sc-RNA seq in intestinal polyp revealed an increased proportion of T_NK cells, and epithelial cells, while plasma cells, mesenchymal cells, and fibroblasts showed a significantly decreased in this patient. GO terms of intestinal polyp-derived fibroblasts suggested that APC/c-MYC signaling modulates fibrotic subpopulation remodeling in intestinal polyp of GS patient. In addition, Sc-RNA seq in scalp cyst revealed an increased proportion in epithelial cells and T cells in this patient. Furthermore, the expression of APC was lowly expressed in endothelial cell, fibroblasts, melanocytes, luminal epithelial, and T cells and the expression of c-MYC was highly expressed in melanocytes, luminal epithelial, and endothelial cell in scalp cyst. Fibroblasts of three tissues was integrated and re-clustered to evaluate the commone menchanisms of fibroblasts remodeling. We identified three fibroblast subpopulations (FC0, FC1 and FC2), the ratio of FC2 was shown a significantly increased in GS patient, and APC-C-MYC signaling might modulates FC2 subpopulation to proliferate fibroblasts in the occurrence of three GS tissues.

**Conclusions:** Using large-scale single cell RNA sequencing, cellular landscape and aetiology-specific alterations were identified in the left atrial dilatation tumor, intestinal polyp and scalp cyst of the proband. APC-C-MYC signaling modulates fibrotic subpopulation remodeling in LAAA and intestinal polyp while epithelial subpopulation remodeling in scalp cyst, indicating that syndrome subtypes caused by the same gene mutation in the same individual may still lead to different cellular and gene expression signatures and heterogeneity. This new approach provides a wealth of novel insights into the molecular changes that underlie the cellular processes relevant for cardiac biology and pathophysiology and also shed light on strategies for cell type- and stage-specific intervention in cardiac diseases.

## 1. Introduction

Gardner’s syndrome (GS), first described by Gardner in 1951 [1], clinical subgroup of familial adenomatous polyposis (FAP), is originally characterized by polyps, osteomas, and soft tissue tumors including pithelial cysts, fibroids, and desmoid tumors. The clinical penetrance of the Gardner’s syndrome is variable, represents a multisystemic disease. Symptoms are usually evident by the 20th year of age, but they may present anytime between 2 mo and 70 years. In general most of the polyps in colonic, gastric and small intestinal adenomatous polyps are small (< 5 mm) and therefore, it is misdiagnosed in clinical practice [2, 3]. Desmoid tumours in GS have an incidence between 8-12%, and usually occur in the small bowel mesentery or unbdominal wall [4, 5]. Extra-abdomina desmold tumours in Gardner’s syndrome are rare [5] and also easily misdiagnosed. Current treatment options for GS are poor, partly because it lacks of the exact molecular pathological mechanism.

GS has an autosomal dominant pattern and most cases show familial clustering, one-third of cases occur due to spontaneous mutations. About 70% patients with FAP and GS have APC germline mutations [6]. APC is a tumor suppressor gene, and up-regulation of its expression can inactivate the Wnt pathway during tumorigenesis [7]. Moreover, APC also lead to the upregulation of c-MYC, which is critical for regulating many biological processes including cell proliferation and apoptosis in some cancers associated with APC mutations [8] However, APC-C-MYC signaling modulating Gardner’s syndrome remain to be elucidated.

Typically, cardiac fibroma is benign neoplasms of the heart that occur in the aortic or mitral valves [9]. Unlike other cardiac fibroma, cardiac fibroma originating from the left atrium is extremely rare, accounting for 1% of all benign cardiac tumours in adults [9]. Given GS present as multi-organ system disease and previous studies mainly focus on the mechanism of a certain organ [10], little is known about the heterogeneity and connection at multi-organ level. In addition, GS was easily misdiagnosed based on small polyps in multisystemic tissue and current effective treatment options to prevent or treat GS are poor, partly because it lacks of the exact molecular pathological mechanism. The identification of precise molecular and cellular targets for GS therapeutic development requires a comprehensive understanding of the cell type-specific responses and cellular heterogeneity in GS.

ScRNAseq was first reported by Collins and Varmus in 2015 in the NEJM [11], and this breakthrough brought novel insight into diseases with identification the heterogeneity of phenotypes within individual cell subtype populations. Previous studies with scRNA-seq on human samples did not attempt to characterize the heterogeneity and connections among multiple organs in a same individual [12]. This present study proves interesting clinical case, as these are limited cases of GS asassociated with left atrial appendage aneurysm using sc-RNA sequencing. Herein, the patient was diagnosed with colonic polyps, multiple osteomas in skull, a giant left atrial appendage fibroma, and thyroid malignancy. Importantly, mutations in APC were identified supporting a diagnosis of Gardner syndrome, and the patient and his patients was found a mutation at exon 15 of the APC gene in this study, thus suggesting its familiar Gardner’s syndrome. We built a single-cell atlas of LAAA, scalp cyst and intestinal polyp in the same GS patient, and explored the heterogeneity and connections among multiple organs and the characteristics and key regulatory pathways of distinct cell subtypes. To the best of our knowledge, this is the first scRNA-seq study that identified pathological mechanism as the cause of GS. These findings will help us understand GS pathogenesis in depth, and provide potential targets for clinical therapies of these diseases.

## 2. Materials and Methods

### 2.1 WES and bioinformatics analysis

To identify the candidate genes for Gardner’s syndrome, we applied WES to search for potential genetic variants in a family with Gardner’s syndrome. WES was performed on three members of the family. The normal human population database consisted of the Thousand Genomes Project (http://browser.1000genomes.org), ESP6500SI-V2 (http://evs.gs.washington.edu/EVS), ExAC Human Exome Integration Database (http://exac.broadinstitute.org/), and the sequencing company’s internal database. All variants were annotated with ANNOVAR software (version 2014). Patients had signed an informed consent form before enrollment. This study was approved by the ethics committee of the Sichuan Academy of Medical Sciences and the Sichuan Provincial People’s Hospital.

### 2.2 Single-cell RNA sequencing

Single-cell sequencing for the clinical samples of left atrium, intestinal polyps and epidermoid cysts was previously described [13]. Briefly, single cells were captured in droplet emulsions using the GemCode Single-Cell Instrument (10x Genomics, Pleasanton, California, USA), and scRNA-seq libraries were constructed as per the 10x Genomics protocol using GemCode Single-Cell 3′ Gel Bead and Library V3 Kit (10x Genomics, Pleasanton, California, USA). The samples were diluted in PBS with 2% FBS to a concentration of 1 × 10^4^ cells per µl. Cells were loaded in each channel with a target output of 1 × 10^5^ cells per sample. All reactions were performed in the BioRad C1000 Touch Thermal cycler with 96-Deep Well Reaction Module. Amplified cDNA and final libraries were evaluated on a Fragment Analyzer using a High Sensitivity NGS Analysis Kit (Advanced Analytical). RNA was controlled for sufficient quality on an Agilent 2100 Bioanalyzer system (Santa Clara, CA, USA) and quantified using a Qubit Fluorometer (Waltham, MA, USA). Libraries were subsequently sequenced on the NovaSeq 6000 Sequencing System (Illumina, San Diego, CA. USA). Raw sequencing data were demultiplexed, aligned, and counted with Cell Ranger pipelines.

### 2.3 Single-cell Analysis

Cellranger mkfastq command was used to generate fastq files, which were later leveraged by the cellranger count command in order to produce expression data at a single-cell resolution. Cellranger aggr command combined sequencing data from multiple libraries with mapped sequencing depth. Pandas (v.1.1.2), NumPy (v.0.25.2), Anndata (v.0.6.19), Scanpy (v.1.8), and Python (v.3.7) were used for pooling multi-sample single-cell counts and for downstream analyses. Total cells were filtered for counts (500 < n_counts < 50000), genes (300 < n_genes < 10000), mitochondrial genes (percent_mito < 20%), and ribosomal genes (percent_ribo < 20%). After filtering, gene expression for each cell was normalized (normalize_per_cell: counts_ per_cell_after = 10,000) and log-transformed (log1p). Highly variable genes were detected (detect(highly_variable_genes)) and PCA was performed based on highly variable genes (top3000). Umap dimensionality reduction and Leiden clustering (resolution 0.1 – 1.2) were carried out, and cell lineages were annotated on the basis of algorithmically defined marker gene expression for each cluster (sc.tl.rank_genes_groups, method=‘wilcoxon’). To aid with the annotation of the subpopulations of each cell compartment, we calculated the differentially expressed genes using the Wilcoxon Rank Sum test with Bonferroni–Hochberg adjustment as it was implemented in the scanpy workflow (log_2_FC > 1 and adjust p-value < 1e-05). Enrichment analyses of these genes were performed using the gene ontology enrichment tool and the Kyoto Encyclopedia of Genes and Genomes (KEGG) pathway analysis using Enrichr [14].

For pseudotemporal analysis, the normalized data from the indicated clusters calculated in Seurat was passed directly into Monocle2 (version 2.6.3)[15]. For gene regulatory network analysis, we generated co-expression networks via arboretum python software libraries (https://github.com/tmoerman/arboretum) and implemented GRNBoost2 [16]. Utilized the SCENIC package (version 0.1.7) to generate cell regulatory networks from our single-cell RNA sequencing data [17].

### 2.4 RNA-seq experiments and data analysis

Total RNA was isolated from all heart samples using a TRIzol reagent (Invitrogen, catalog no. 15596018) prepared into cDNA library according to standard Illumina protocols. Libraries were sequenced using 150 bp paired-end sequencing on Illumina Novaseq platform. Raw reads of fastq format were firstly processed through in-house perl scripts, clean reads were obtained by removing reads in low quality and adaptor contamination, and Q20, Q30 and GC content were calculated in the clean data. The clean reads were further mapped to Ensemble mouse genome (mm10) using HISAT2 with default parameters. Only uniquely mapped reads were counted for gene expression. Differential expression analysis was performed using the DESeq2 R package (1.16.1). The resulting P-values were adjusted using the Benjamini and Hochberg’s approach for controlling the false discovery rate. Genes with an adjusted P-value <0.05 found by DESeq2 were assigned as differentially expressed. We use the local version of Gene Set.Biological process (BP) of Gene Ontology was used for GSEA independently. The heatmap and bubble plot was generated using on online platform (http://www.bioinformatics.com.cn).

### 2.5 Immunostaining

Tissue samples were fixed with 4% (vol/vol) paraformaldehyde for 15 min and then permeated and blocked with 0.3% (vol/vol) Triton X-100 and for 5% BSA in goat serum solution for 1 h at room temperature. The samples were then incubated with primary antibody against CD34 (1:400) and beta-catenin (1:400) overnight at 4 °C. Next day, the samples was incubated with secondary fluoresce-labeled anti-mouse/rabbit antibody (1:500) for 1 h at room temperature. The morphology of the tissue was analyzed by hematoxylin and eosin (HE) staining. Fibrosis was analyzed by Sirius Red staining. Images were captured under Leica sp8 confocal microscopy.

### 2.6 Statistics and reproducibility

All results are expressed as means ± SEM. Student’s t-test was used for comparison of two groups, one-way ANOVA, post hoc or two-way ANOVA was used for comparison of three groups in the manuscript, *p < 0.05, **p < 0.01, ***p < 0.001, n.s., not significant. p < 0.05 was considered statistically significant. All representative images or results were selected from at least three independent experiments with similar results.

## 3 Results

### 3.1 Cellular landscape and aetiology-specific alterations of left atrium in GS patient

A man in his 30s was experiencing episodes of palpitations, dyspnea, and fatigue, which is short-lasting and irregular in nature and these symptoms had begun 4 months earlier, The patient had a negative medical history, and without history of diabetes mellitus, hypertension and familiar cardiovascular diseases.

Here, we found a familiar Gardner syndrome combined left atrial appendage fibroma, which was rare in patients with GS. To investigate first how LAAA alters the molecular and cellular profiles, we performed a transcriptome analysis of left atrium tissue from this patient by scRNA-seq. We sampled 12,805 cells from LAAA. A total of 12,277 cells were obtained after quality control. For the identification and classification of cell populations from both LAAA, uniform manifold approximation and projection (UMAP) were used and divided cells into six major cell types according to known cell-type markers, including cardiomyocyte (TNNT2+) cells (CMCs), endothelial (VWF+) cells (ECs), fibroblast (COL1A1+) cells (FBs), macrophage (CD163+) cells (MCs), smooth muscle (MYH11+) cells (SMCs), and T (IL7R+) cells (TCs) in in Figure 1A and 1B. These cell subsets are identified by some of marker genes in Figure 1C and 1D. These results reveal the cell type diversity in diseased LA tissue.

**Figure 1.**
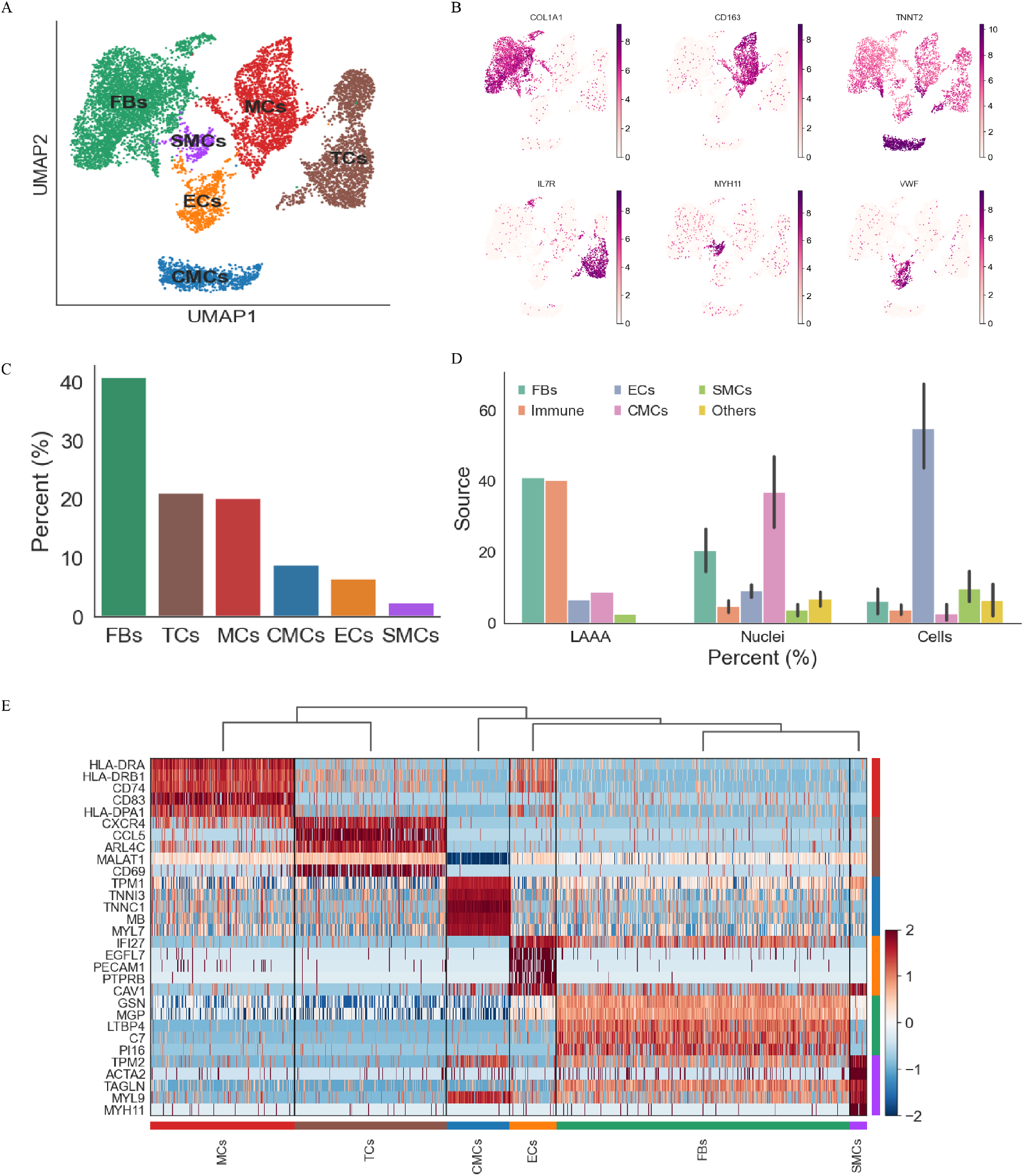
The UMAP plots of single-Cell transcriptomic sequencing, clustering of cells profiles in left atrium of GS patient. (A) Left atrium cells were assigned cells into 6 different groups. (B) Marker genes were shown in 6 different groups. (C) The proportions of cell types were analyzed in 6 different groups. (D) The proportions of cell types were compared between this study and previous study. (E) The top five marker genes were shown in 6 different groups.

Interestingly, comparing with normal healthy LA tissue in previous study[21], the most striking feature of these cells is a increased ratio of fibroblasts and immune cells revealed by LAAA patient in Figure 1E. A characteristic feature of some cancers is an excessive tumor microenvironment with infiltrated immune cells that include macrophages (TAMs), and high numbers of activated fibroblasts [22], also known as myofibroblasts. These results suggested diseased LAAA with GS patient appears to display tumor-prone properties.

### 3.2 Remodeling of fibroblasts populations in GS patient with LAAA

As an increased proportion of fibroblasts were observed in GS patient with LAAA, consistent with the previous clinical phenotype, the whole population of fibroblasts was again re-clustered to further evaluate the effects of LAAA on remodeling of fibroblasts populations (Figure 2A and B). We re-clustered fibroblasts and identified five distinct sub-populations: cancer associcated fibroblasts (CAF, FOSB+), cardiomyocyte like fibroblasts (CLF, TNNT2+), endothelial like fibroblasts (ELF, PTGDS+), T_cells like fibroblasts (TLF, CCL5+), and nomal like fibroblasts (NLF, LAMA2+) in Figure 2A and 2B. Differences between five distinct sub-populations could also be identified by expression of some marker genes in Figure 2B and 2C. Notably, CAF constituted the main component of cardiac fibroblasts in LAAA, indicating that NLF in tumor region was reprogrammed into CAF, which has been previously reported in some cancers [23].

**Figure 2.**
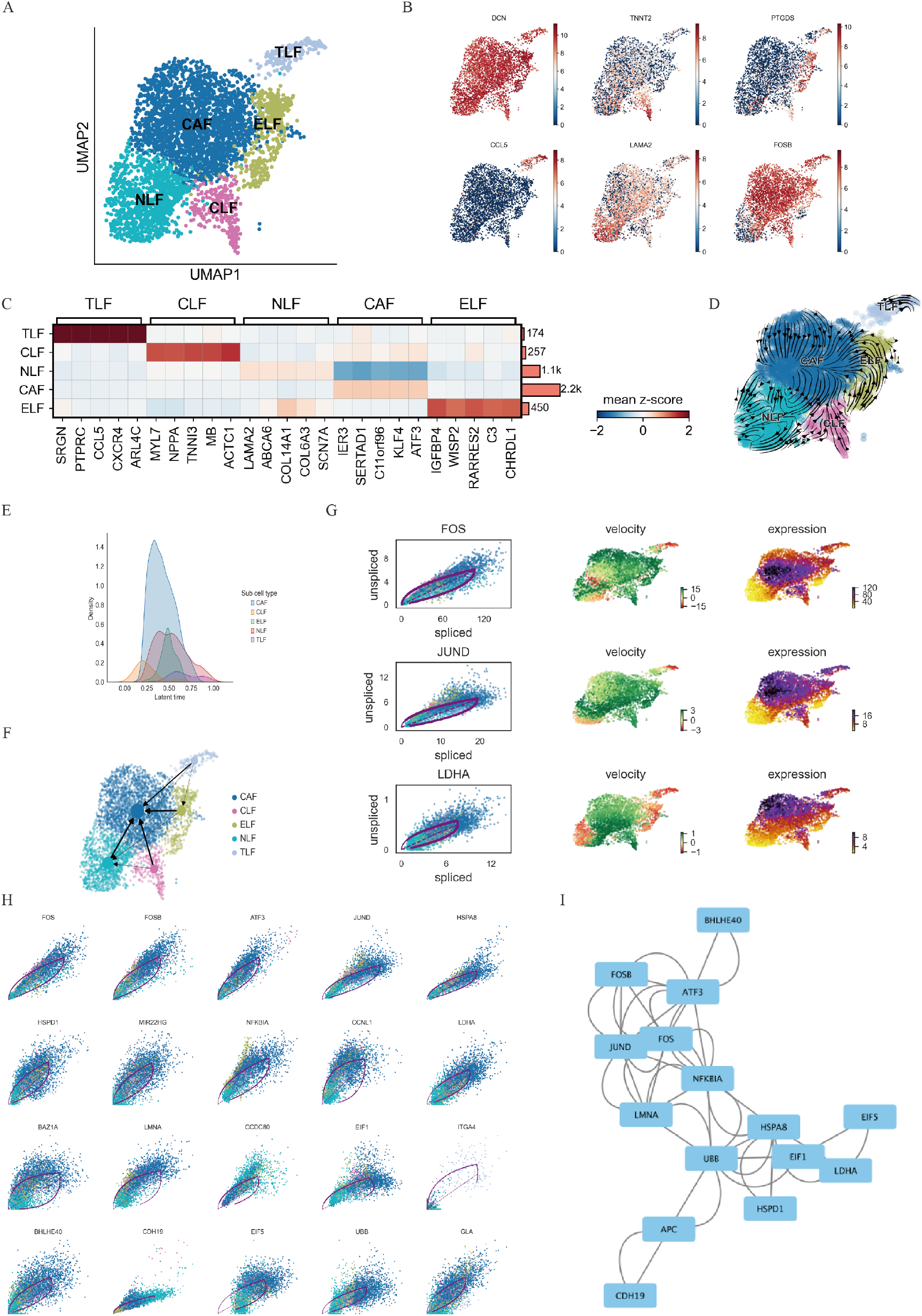
Remodeling of fibroblasts populations in GS patient with LAAA. (A) LA fibroblasts were assigned into five subgroups. (B) An important marker gene was shown in each subgroup. (C) Marker genes were shown in two subgroups in LA fibroblasts. (D) UMAP plotting RNA velocity analysis of subclustered fibroblasts undergoing state transitions. (E) Latent time distribution of trajectory-associated fibroblast clusters. (F) Pseudotime analysis of the subgroups in left atrium of GS patient. (G) The dominant CAF state was characterized by expression of FOS, JUND and LDHA. (H) The 20 most important genes were selected in pseudotime analysis.

To further investigate this ongoing process, we performed trajectory analysis and RNA velocity analysis on narmal fibroblasts and CAF. Generally, fibroblast cells are thought to be non-dividing, terminally differentiated [23], but an increase in velocity length indicates they are capable of changing phenotypic state once cycled along a lineage. Interestringly, fibroblasts from LAAA have unique maturation pathways that culminate in one distinct terminal states: cancer associcated fibroblasts (CAF). LAAA fibroblasts made a preferential transition from the immature phase (CLF, ELF, TLF, and NLF) at the apex of the trajectory to mature phase of tumor-like properties (*FOSB-*expressing; CAF) in Figure 2B and 2D. PAGA graphs reveal the denoised topology of the data at a chosen resolution and collectively transitioned from four fibrotic subgroups to CAF (Figure 2F). These results further indicated that normal fibroblast was reprogrammed into CAF in LAAA. Combined with molecular features, we recognized that upregulation of tumor-specific genes FOS, JUND and LDHA might lead to the dominant CAF state (Figure 2G), which may explain the increased fibroblasts by immunohistochemistry and single-cell sequencing. Together, these results showed that LAAA fibroblasts have dynamic programming abilities that result in high fibroblast proliferation ability drived by CAF states in LAAA patient with GS.

To understand how APC regulates the process of dynamic fibroblast programming, PPI network was constructed to depict the complex regulatory pathways. The 20 most important genes were selected in pseudotime analysis (Figure 2H). In addition, we noticed that 18 of 20 genes (FOS, JUND, LDHA, ect.) were all upregulated in this differentiation process, while CDCC80 and were downregulated (Figure 2H). Similar phenomenon was observed in PPI network, where APC interacted with 11 of these 18 upregulated genes on the side, while APC interacted with CDH19 on the other side, indicating that these two types of genes may have different functions in the development of LAAA (Figure 2I). These results provide potential targets for inhibiting or reversing the formation of CAF in LAAA. All these data indicated that APC-FOS-JUN signaling modulates fibrotic subpopulation remodeling in LAAA.

### 3.3 Cellular landscape and aetiology-specific alterations of intestinal polyp in GS patient

To explore the impact of intestinal polyp (IP) on cellular landscape, we performed scRNA-seq on 20,381 cells, which were isolated from intestinal tissue from a healthy site and intestinal polyp. A total of 15,460 cells were obtained after quality control.

We further clustered cells across all samples according to their transcriptional profiles (Figure 3A). Reclustering of the cellular composition from intestinal tissue identified eight major cell types: endothelial cells (e.g., PECAM1+, EGFL7+, CLDN5+), epithelial cells (e.g., KRT8+, KRT18+, ELF3+), fibroblasts (e.g., CXCL14+, CXCL3A1+, COL6A2+), mesenchymal cells (e.g., NRXN1+, CRYAB+, CLU+), plasma cells (e.g., JCHAIN+, MZB1+, IGHA1+), mast cells (e.g., TPSAB1+, TPSB2+, CPA3+), B cells (e.g., HLA_DRA+, HLA_DPB1+, CD74+), and T_NK cells (e.g., PTPRC+, IL32+, CCL5+) in Figure 3A and 3B.

**Figure 3.**
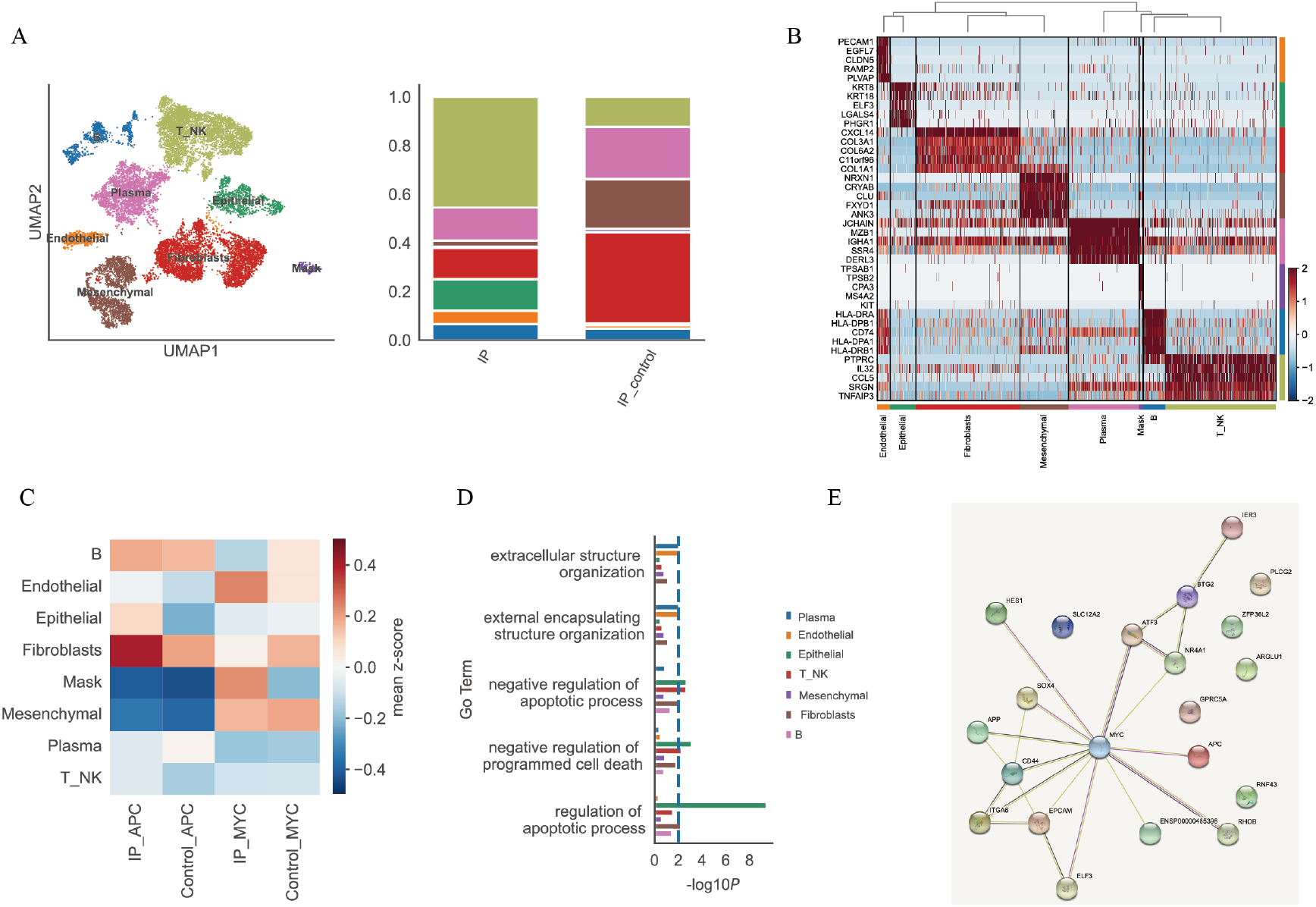
The UMAP plots of single-Cell transcriptomic sequencing, clustering of cells profiles in intestinal polyp of GS patient. (A) The cells of intestinal polyp were assigned cells into eight different groups. The proportions of cell types were further analysed in gut tissues. (B) Gene expression profile of marker genes in eight gut subgroups. (C) The expression of *APC* and *c-MYC* was shown in eight intestinal polyp subgroups. (D) The enriched biological process of intestinal polyp subgroups. (E) The complex regulatory pathway may be regulated by APC dysfunction. IP: intestinal polyp; IP_control: healthy intestinal tissue.

The distributions of cell types were further analysed in intestinal polyp from a healthy individual and this patient. The distributions of cell types in intestinal polyp (GS vs healthy individuals) contain endothelial cells (6% vs 1%), epithelial cells (10% vs 2%), fibroblasts (10% vs 36%), B cells (6% vs 5%), plasma cells (12% vs 22%), mesenchymal cells (1% vs 20%), mast cells (1% vs 3%), and T_NK cells (45%vs 10%) in Figure 5A. Interestingly, compared with a healthy individual, the ratio of T_NK cells, epithelial cells, and epithelial cells revealed a significantly increased in GS patient, while plasma cells, mesenchymal cells, and fibroblasts showed a significantly decreased in GS patient.

**Figure 4.**
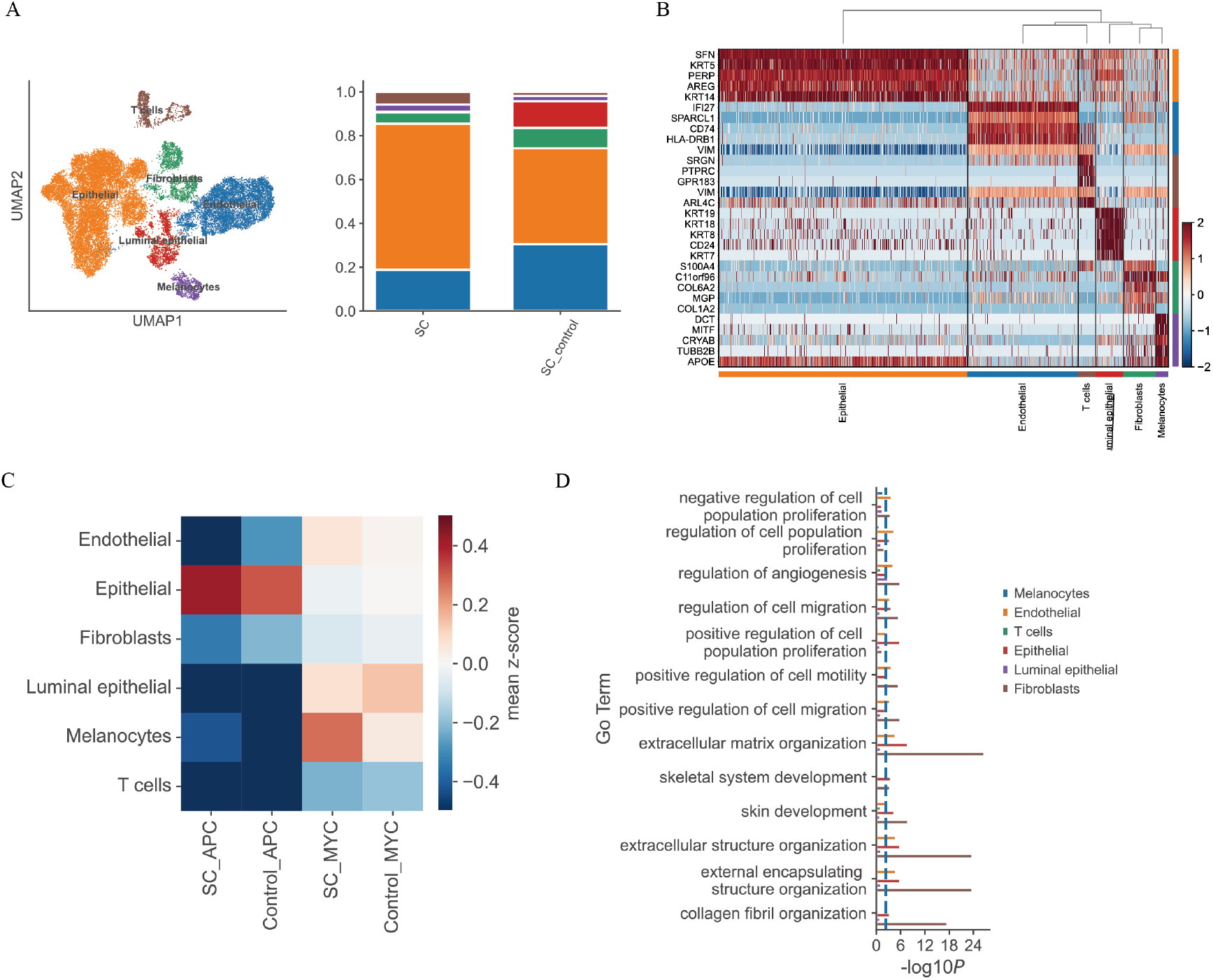
The UMAP plots of single-Cell transcriptomic sequencing, clustering of cells profiles in scalp cyst of GS patient. (A) The cells of scalp cyst were assigned cells into 6 different groups. The proportions of cell types were further analysed in head tissues. (B) Gene expression profile of marker genes in six head subgroups. (C) The expression of *APC* and *c-MYC* was shown in eight scalp cyst subgroups. (D) The enriched biological process of scalp cyst subgroups. SC: scalp cyst; SC_control: healthy scalp tissue.

**Figure 5.**
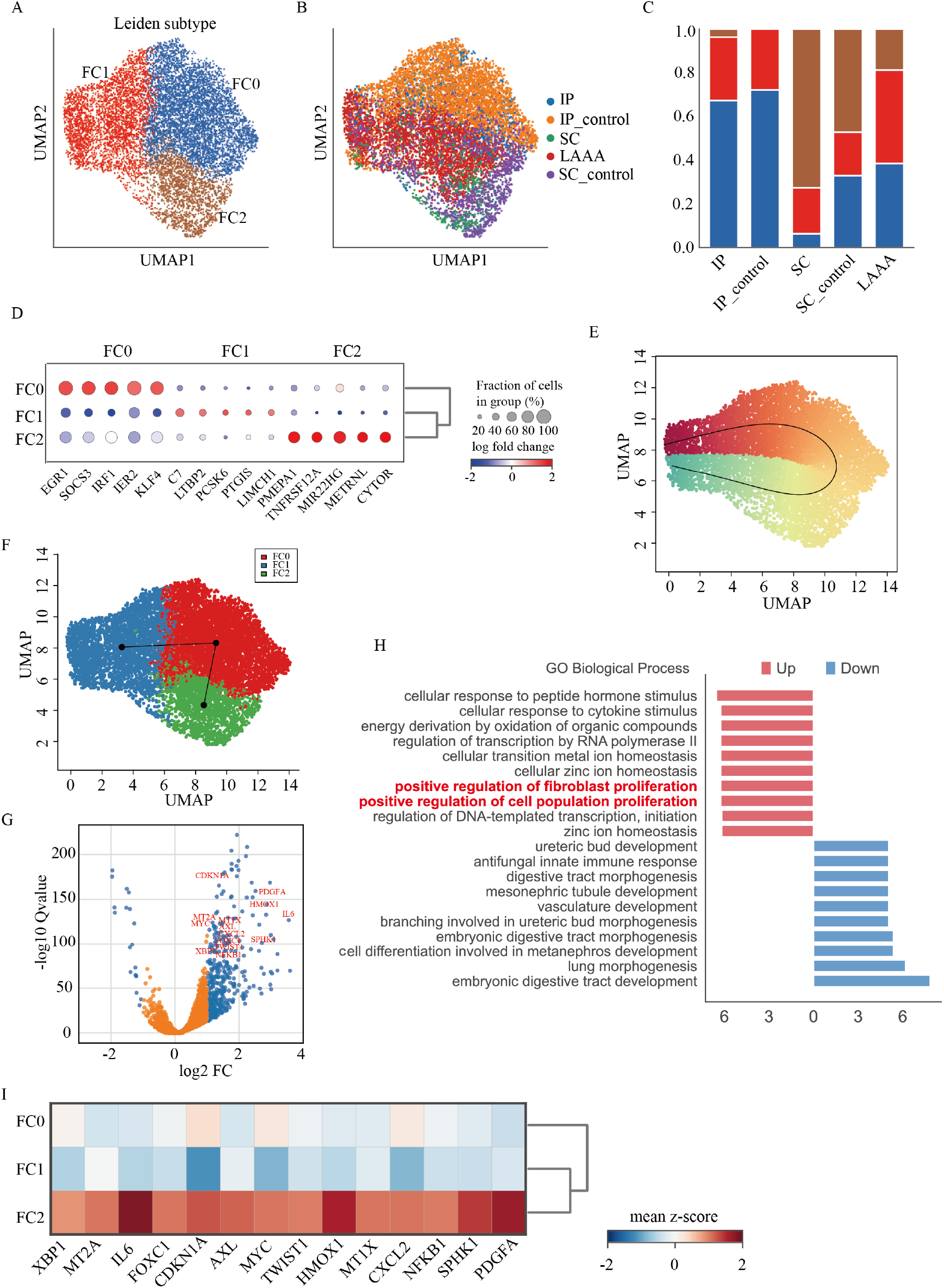
Remodeling of fibroblasts populations in three tissues with GS patient. (A) The fibroblasts of three tissues (intestinal polyp, scalp cyst, and LAAA) were assigned cells into three different groups. (B) The fibroblasts of three tissues were sourced from these GS patient and healthy control. IP: intestinal polyp; IP_control: healthy intestinal tissue; SC: scalp cyst; SC_control: healthy scalp tissue; LAAA: left atrial appendage aneurysm. (C) The proportions of cell subtypes were further analysed in three tissues. (D) Gene expression profiles of marker genes in three fibroblast subgroups. (E) Pseudotime analysis of fibroblasts. (F) Pseudotime analysis of three fibroblast subgroups. (G) Volcano plots analysis of differential expression genes between FC2 subgroups and others. (H) The enriched biological process of FC2 subgroups compared with others. (I) Gene expression profiles of fibroblast and cell population proliferation upregulated in FC2.

Furthermore, the expression of APC was lowly expressed in B cells, epithelial cells, fibroblasts and the expression of c-MYC was highly expressed in endothelial cells, fibroblasts, plasma cells, epithelial cells, and T_NK cells of GS patient (Figure 3C). Interestingly, there were 123 up-regulated genes in intestinal polyp-derived fibroblasts, involving with GO terms including regulation of apoptotic processs, programmed cell death, and lymphocyte migration (Figure 3D). The complex regulatory pathway may be regulated by APC dysfunction (Figure 3E). APC also lead to the upregulation of c-MYC, which is critical for regulating many biological processes including cell proliferation and apoptosis in some cancers associated with APC mutations [8]. These results suggested that APC/c-MYC signaling modulates fibrotic subpopulation remodeling in intestinal polyp of GS patient.

### 3.4 Cellular landscape and aetiology-specific alterations of scalp cyst in GS patient

To understand the impact of scalp cyst (SC) on cellular landscape, we performed scRNA-seq on 26,237 cells, which were isolated from scalp tissue from a healthy site and this patient with GS. A total of 24,262 cells were obtained after quality control.

Next, we clustered cells across all samples according to their transcriptional profiles (Figure 4A). Reclustering of the cellular composition from scalp tissue identified seven cell types: epithelial cells (e.g., SFN+, KRT5+, PERP+), endothelial cells (e.g., IFI27+, SPARCL1+, CD74+), fibroblasts (e.g., S100A4+, C11orf96+, COL6A2+), melanocytes (e.g., DCT+, MITF+, CRYAB+), luminal epithelial (e.g., KRT19+, KRT18+, KRT8+), and T cells (e.g., SRGN+, PTPRC+, GPR183+) (Figure 4A and 4B).

The proportions of these cell types were next analysed in scalp tissues from normal scalp tissues and scalp cyst. The cell types of scalp cyst and normal scalp tissues showed distinct relative cell number ratios (Figure 4A). Interestingly, Increased proportions were observed for epithelial cells and T cells in scalp cyst of these GS patient, which is consistent with reports that scalp cyst exhibit increased epithelial cells (Figure 4A). The proportions of luminal epithelial, fibroblasts, and endothelial cells were decreased in scalp cyst tissues compared to normal scalp tissues, perhaps resulting from the excessive expansion of epithelial cells and T cells.

Furthermore, the expression of APC was lowly expressed in endothelial cell, fibroblasts, melanocytes, luminal epithelial, and T cells and the expression of c-MYC was highly expressed in melanocytes, luminal epithelial, and endothelial cell of GS patient (Figure 4C). Interestingly, differentially regulated genes in scalp cyst, involving with GO terms including regulation of cell population proliferation, regulation of angiogenesis, and cell migration (Figure 4D).

### 3.5 Commone menchanisms of fibroblasts remodeling in three tissues with GS patient

To understand subcellular landscape and aetiology-specific alterations of fibroblasts in LAAA, scalp cyst, and intestinal polyp, the whole population of fibroblasts was integrated and re-clustered to evaluate the effects of three pathological tissues on remodeling of fibroblasts (Figure 5A). There were three fibroblast subpopulations, including FC0, FC1 and FC2. These cell subsets are identified by some of marker genes in Figure 5B. Interestingly, the ratio of FC2 was shown a significantly increased in GS patient, compared with healthy control, indicating that FC2 might play an important role in the occurrence and development of GS.

To discover the relationship between the various subgroups, we performed a pseudotime analysis using the Slingshot package. Pseudotime analysis found that the three cells (FC0, FC1 and FC2) have a close evolutionary relationship (Figure 5C). PAGA graphs reveal the denoised topology of the data at a chosen resolution and reveal its connected and disconnected regions (Figure 5D). These results reveal fibroblast subpopulations FC2 are terminally differentiated cell subsets (Figure 5C and 5D), further indicating FC2 might play an important role in GS.

Next, we investigated underlying mechanism of subpopulation FC2 in GS, and differential expression genes (DEGs) were found using volcanic map in FC2 fibroblasts compared with others. GO enrichment analysis revealed that these DEGs were enriched in positive regulation of fibroblast and cell population proliferation (Figure 5E). There were 14 DEGs of FC2 fibroblasts in these two processes of fibroblast and cell population proliferation, and these DEGs were upregulated in FC2 fibroblasts (Figure 5F). In addition, the expression of MYC was relatively high expressed in FC2, while its expression was lowly expressed in FC0 and FC1. These results provide potential targets for inhibiting or reversing the formation of FC2 in GS. Together, these data indicated that fibrotic remodeling of FC2 subpopulation might continue to proliferate fibroblasts in the occurrence and development of GS.

## 4 Discussion

Although Gardner’s syndrome (GS) has been extensively studied, key mechanisms underlying these diseases are still far from elucidated. GS, a subtype of familial adenomatous polyposis (FAP), represents a rare (1:1 000 000) multisystemic disease [2, 24]. Three features of polyps, osteomas, and soft tissue tumors (epithelial cysts, fibroids, desmoid tumors) were originally described as GS [25]. Given GS present as multi-organ system disease and previous studies mainly focus on the mechanism of a certain organ [10], little is known about the heterogeneity and connection at multi-organ level.

Herein, the patient was diagnosed with colonic polyps, scalp cyst, a giant left atrial appendage fibroma, and thyroid malignancy. Importantly, mutations in APC were identified supporting a diagnosis of GS, and the patient and his patient was found a mutation at exon 15 of the APC gene in this study, thus suggesting its familiar GS. The identification of precise molecular and cellular targets for GS therapeutic development requires a comprehensive understanding of the cell type-specific responses and cellular heterogeneity in GS. We built a single-cell atlas of left atrial appendage fibroma, scalp cyst and intestinal polyp in the same GS patient, and explored the heterogeneity and connections among multiple organs and the characteristics and key regulatory pathways of distinct cell subtypes. To the best of our knowledge, this is the first scRNA-seq study that identified pathological mechanism as the cause of GS. These findings will help us understand GS pathogenesis in depth, and provide potential targets for clinical therapies of these diseases.

Our scRNA-seq analysis identified cellular landscape and aetiology-specific alterations of the left atrial dilatation tumor, and the most striking feature of these cells was an increased ratio of fibroblasts and immune cells in LAAA with GS patient. A characteristic feature of some cancers is an excessive tumor microenvironment with infiltrated immune cells that include macrophages (TAMs), and high numbers of activated fibroblasts. These results suggested diseased LA tissue with GS patient appears to display tumor-prone properties.

Interestingly, We re-clustered and identified five distinct fibroblast subgroups in LAAA: cancer associcated fibroblasts (CAF), cardiomyocyte like fibroblasts (CLF), endothelial like fibroblasts (ELF), T_cells like fibroblasts (TLF), and nomal like fibroblasts (NLF). Notably, NLF should constitute the main component of normal cardiac fibroblasts in LA tissues, while CAF mainly dominated in LAAA tissues, indicating that NLF in tumor region was reprogrammed into CAF, which has been previously reported in some cancers [23]. The trajectory and RNA velocity analysis further confirmed that LAAA fibroblasts made a preferential transition from the immature phase (CLF, ELF, TLF, and NLF) at the apex of the trajectory to mature phase of tumor-like properties (CAF). In contrast to normal fibroblasts, CAFs has been shown to increase autocrine signaling ability and proliferation tendencies in multiple tumor types. According to molecular features, we recognized that upregulation of tumor-specific genes FOS, JUND and LDHA might lead to the dominant CAF state. PPI networks further indicated that APC-FOS-JUN signaling modulates fibrotic subpopulation remodeling in LAAA. These data showed that LAAA fibroblasts have dynamic programming abilities that result in high fibroblast proliferation ability drived by CAF states in LAAA patient with GS. Together, these results suggested diseased LA tissue with GS patient appears to display tumor-prone properties and provide potential targets for inhibiting or reversing the formation of CAF in LAAA.

Our scRNA-seq analysis identified seven cell types: epithelial cells, endothelial cells, fibroblasts, melanocytes, and luminal epithelial. We found aetiology-specific cellular alterations of scalp cyst in GS patient: increased proportions of epithelial cells and T cells and dncreased proportions of luminal epithelial, fibroblasts, and endothelial cells. We re-clustered and identified three scalp epithelial populations: EP1, EP2, and EP3. We noticed that EP1, EP2, and EP3 were composed of epithelial cells from normal scalp tissue, while EP3 of epithelial cells were absent in scalp cyst of GS patient, perhaps resulting from the excessive expansion of EP1 and EP2 in scalp cyst. GO enrichment analysis further revealed that Wnt signaling pathway were upregulated in EP1, and epidermal cell and keratinocyte differentiation, and smooth muscle cell proliferation were enriched in EP2. Generally, tumor suppressor gene APC could inhibit WNT signaling pathway, whlie mutational inactivation of APC leads to hyperactivation of the Wnt pathway [26]. Wnt signaling pathway was activated in EP1 and a mutation of the APC gene was found in these GS patient, which is consistent with reports [6]. These results indicated that APC might regulate Wnt signaling pathway to cell proliferation of EP1 in the development of scalp cyst on GS patient. In addition, we analyzed and re-clustered fibroblasts and identified two distinct sub-populations (FB1 and FB2), and there is no difference in the proportion of fibroblasts sub-populations compared with normal tissue in scalp cyst, which is different from LAAA. These results indicted that the heterogeneity among multiple organs in same GS patient.

We further explore the impact of intestinal polyp on cellular landscape, and reclustered of the cellular composition from intestinal tissue identified eight major cell types: endothelial cells, epithelial cells, fibroblasts, mesenchymal cells, plasma cells, mast cells, B cells, and T_NK cells. Interestingly, the ratio of T_NK cells, epithelial cells, and epithelial cells revealed a significantly increased in GS patient, while plasma cells, mesenchymal cells, and fibroblasts showed a significantly decreased in GS patient. Furthermore, we noticed that the expression of APC was lowly expressed in B cells, epithelial cells, fibroblasts and the expression of c-MYC was highly expressed in endothelial cells, fibroblasts, plasma cells, epithelial cells, and T_NK cells of GS patient. Interestingly, GO terms revealed that regulation of apoptotic processs, programmed cell death, and lymphocyte migration was enriched in intestinal polyp-derived fibroblasts. The complex regulatory pathway further found that APC/c-MYC signaling modulates fibrotic subpopulation remodeling in intestinal polyp of GS patient.

The whole population of fibroblasts were integrated and re-clustered to evaluate the effects of three pathological tissues on remodeling of fibroblasts and connection at multi-organ level. There were three fibroblast subpopulations, including FC0, FC1 and FC2. Interestingly, the ratio of FC2 was shown a significantly increased in GS patient, compared with healthy control, indicating that FC2 might play an important role in the occurrence of GS. Pseudotime analysis found that fibroblast subpopulations FC2 are terminally differentiated cell subsets; further indicating FC2 might play an important role in GS. GO enrichment analysis revealed that these DEGs of FC2 were enriched in positive regulation of fibroblast and cell population proliferation. In addition, the expression of MYC was relatively high expressed in FC2, while its expression was lowly expressed in FC0 and FC1. These results provide potential targets for inhibiting or reversing the formation of FC2 in GS. Together, these data indicated that fibrotic remodeling of FC2 subpopulation might continue to proliferate fibroblasts in the occurrence and development of GS.

In summary, our transcriptome profiling results characterize the heterogeneity and connections among multiple organs in a same GS patient. These results provide a useful resource for understanding the pathological roles of fibrotic cells and APC/c-MYC signaling modulates fibrotic subpopulation remodeling in Gardner’s syndrome. Furthermore, our data can aid the identification of fibrotic molecular targets for the therapeutic restoration and the amelioration of the pathological progression of GS.

## Data Availability

All data produced in the present study are available upon reasonable request to the authors.

## Acknowledgements

The authors would like to thank all patients who provided samples used in this study.

